# The MU Study of Seropositivity and Risk for SARS-CoV-2 and COVID-19: Crucial Behavioral and Immunological Data from Midwestern College Students

**DOI:** 10.1101/2022.01.24.22269758

**Authors:** Tyler W. Myroniuk, Joan M. Hermsen, Christal Hamilton, Ifeolu David, Michelle Teti, Yerina S. Ranjit, Shannen N. Woodrey, Julie A.W. Stilley, Emma Teixeiro, Yue Guan, Mark Daniels, Enid Schatz

## Abstract

**Objective:** We describe our Fall 2020 study of college students’ COVID-19 related behaviors, attitudes, and antibody test results.

**Participants:** The study included 1,446 randomly selected and self-enrolled undergraduate and graduate students from a midwestern university.

**Methods:** An online survey was distributed to a sample of students, between September and December 2020. A sub-group also participated in a SARS-CoV-2 antibody blood draw.

**Results:** Nearly half of students reported a prior COVID-19 test with 22% indicating a positive test, which represents an 11% positivity rate across all student participants. Of those who participated in antibody testing, 15.1% tested positive for SARS-CoV-2 antibodies. Seventy-seven percent of participants said they would get vaccinated. One-third of students reported moderate to severe generalized anxiety disorder and 13% reported moderate to severe depression.

**Conclusions:** This study informed campus decisions in Fall 2020. The importance of effective public health messaging on campus should continue in the future.

## 1 Introducing Our Transdisciplinary Study

In the Fall 2020 semester, our transdisciplinary research team collected data on seropositivity of, and behaviors related to, SARS-CoV-2 among students, faculty, and staff at a large midwestern university (MU). Our team’s mission was to understand the social *and* hard science of COVID-19 in a college setting, in efforts to inform campus COVID-19 mitigation strategies. The *MU Study of Seropositivity and Risk for SARS-CoV-2 and COVID-19* went beyond the more-prominent COVID-19 related research among college students by incorporating highly accurate antibody testing to examine trends in SARS-CoV-2 seropositivity on campus.^1,2,3^ The study also included an online survey focused on attitudes and behaviors related to COVID-19 and public health.

This paper outlines our study design and sample, then describes college students’ results from the Fall semester emphasizing four key study variables: seropositive and self-reported Polymerase Chain Reaction (PCR)-positive test rates; self-reported chances of about becoming infected with COVID-19; attitudes towards vaccination; and anxiety and depression.

The collaboration of the study’s primary investigators from various departments including communication, health management and informatics, molecular microbiology and immunology, public health, sociology, and veterinary medicine, formed a natural group devoted to tackling the COVID-19 crisis and informing MU’s Incident Command and human health experts on campus. These efforts resulted in campus public health safety adjustments throughout the Fall 2020 semester.

## 2 Research Methodology and Analytic Sample

### 2.1. Design

The full study, which ran from September through December 2020, included students, faculty, and staff affiliated with the university. The study was originally by invitation only; the MU administration periodically provided stratified, randomly drawn lists of student and employee email addresses for 950 undergraduates, 250 graduate/professional students, and 400 faculty/staff. The first three draws were sent email invitations every 10 days. Low response rates and the need to enroll a higher number of individuals for antibody testing in order to calculate disease prevalence led us to send two sets of 1,600 invitations, weekly, starting on October 12, 2020. In mid-October, we also received MU Institutional Review Board (protocol#2028427) permission to allow individuals to self-enroll into the study. We advertised self-enrollment via campus-wide email distribution lists, Facebook, and other social media, while continuing with the weekly targeted 3,200 email invitations. In the end, nearly 18% of participants came from self-enrollment, adding a crucial supplemental sample to this study.

In total, 28,217 faculty, staff, and students were randomly recruited (9.0% participation rate in at least study element—antibody testing and/or online survey—N=2,530) while 670 self-enrolled (with 83.4% then participating in at least study element). The total participation rate, in at least one study element—antibody testing and/or the online survey—was 10.7% (N=3,089). Out of these 3,089 participants, 1,446 students and 1,511 faculty/staff completed the survey (N=2,957; 95.7%). Of this group who completed the survey, 62.4% of students (N=902) and 77.4% of faculty/staff (N=1,170) also provided a blood sample for antibody testing.

Whether invited or self-enrolled, individuals were sent information via email, and eligible persons were asked to complete the survey and blood draw for antibody testing within two weeks of initial invitation or enrolling. All blood and survey data were collected and stored in <u>REDCap</u>. Those who wished to participate in the blood draw used an online platform created for the project to make an appointment at an on-campus student health facility. Study participants were eligible to participate in a raffle as compensation for their contribution to each study component.

MU investigators and laboratory staff analyzed the 20mL blood samples using a Siemens Healthineers’ testing system designed to identify SARS-CoV-2 antibodies with high precision. This corporate partnership was essential because an internally developed antibody test was not available at MU.

### 2.2. Reasoning

Antibody testing enabled us to track changes in the percent of participants presenting with positive antibodies over the course of the Fall semester. The presence of antibodies is a “historical” view of having been infected with SARS-CoV-2 as they usually appear one to three weeks after infection and may not last more than a few months after infection.^4,5,6^ The antibody tests provided weekly snapshots that correlated with what the likely caseload was in previous weeks.

The survey collected general demographic and socio-economic data, as well as data on mental health, attitudes about COVID-19 mitigation strategies, and perceptions about how well individuals and the community were doing in combating COVID-19.^7^ In the interests of scientific transparency, we present descriptive statistics that include item non-response in the results section.

### 2.3. MU Students in the Study

Among the 1,466 student participants, 62.9% were undergraduates, 30.3% were graduate students or in professional degrees, 0.9% were another type of student, and 5.9% did not provide this information. Of the student participants, 80.8% identified as White, 4.7% as Hispanic/Latino, 4.0% as African, Black or African American, and 9.9% as another racial/ethnic identity, while 0.6% did not respond. Also, 69.2% of participants identified as female, 28.6% as male, and 1.8% as outside of the gender binary.

## 3 Results

### 3.1. Self-reported COVID-19 Testing and Positivity Rate

Table 1 presents the descriptive statistics for key variables from the study. As shown in the second column, nearly half (49.2%) of students had received at least one PCR test since the onset of the pandemic and prior to taking the survey. Of the 708 students who reported having had a PCR test, 22% had at least one positive test (n=159), which represented 11% of all student survey respondents.

**Table 1.**
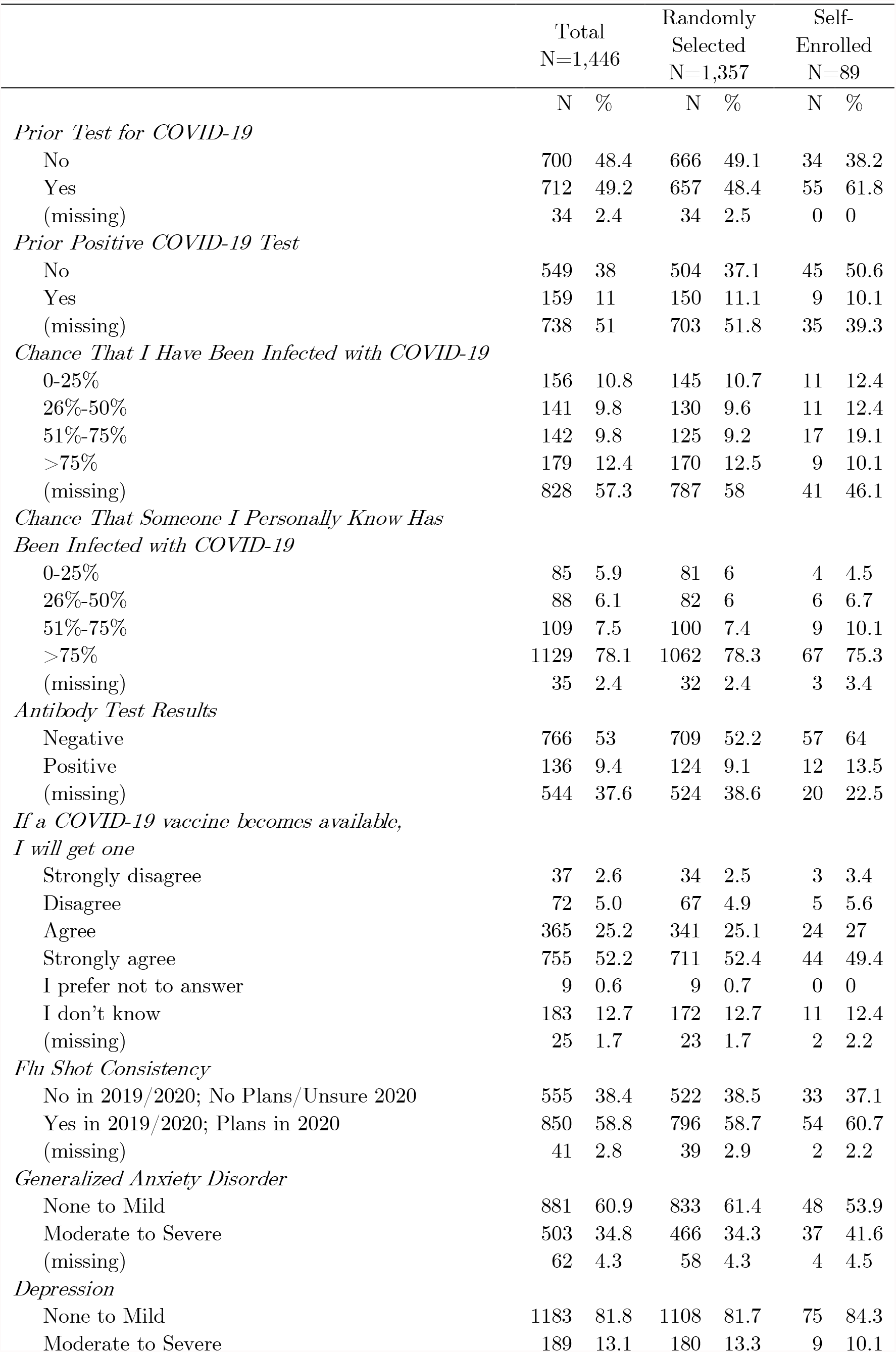
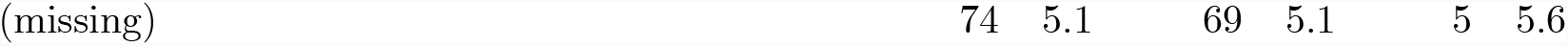
MU Students, Fall 2020 Semester

### 3.2 SARS-CoV-2 Antibodies Testing and Positivity Rate

Only 62% of students participated in the blood draw for antibody testing (N=902). Of those who donated their blood, 15.1% tested positive for SARS-CoV-2 antibodies (authors’ calculations). This positivity rate is higher than the positive PCR self-report rate of 11% among the students in the study. Note, however, that these samples are not directly comparable.

### 3.3 Perception Chance of COVID-19 Infection

Students were asked what the chances (0-100%) were they and someone they personally know had been infected with COVID-19. Over twelve percent of students said there was a 75% chance or greater they had been infected—a figure close to the 11.0% self-report positivity rate found for the student participants. This contrasts with the 78.1% of students who said there was a 75% chance or greater they knew someone personally who had been infected. In other words, while a relatively low share of students reported a high likelihood of COVID-19 infection, a relatively high share indicated a high likelihood they knew someone with COVID-19 infection. This discrepancy between self-report of COVID-19 infection chances and perceptions of risk for others is due in part to the high share of students who did not answer the question on personal chances of COVID-19 infection. Indeed, the majority of students (57%) were unwilling to “guess” if they had been infected with COVID-19. We are unable to discern why this is the case. We suspect it is related to student concerns about how the information would be used to restrict student participation in classes and campus life.

### 3.4. Vaccination Intentions

Students show a high level of vaccine intent. More than three quarters of students (77.4%) agreed or strongly agreed they would get a COVID-19 vaccine if it became available. Considering that herd immunity would require somewhere between 70% to 80% of the US population to become vaccinated, it is possible the campus will reach herd immunity if students follow through on their vaccine intentions.^8,9^ Yet, nearly 1 in 5 students do not plan to get the vaccine or were unsure at the time of the survey. Further, despite continuous public discourse about the benefits of vaccines throughout the pandemic, only 58.8% of students indicated that they had their flu shot in 2019 and had already received or planned to get their shot in 2020. This rate is remarkably low and does not bode well for future campus infectious disease mitigation strategies for Fall 2021 and beyond.

### 3.5 Mental Health Outcomes

Lastly, our externally validated mental health indicators of General Anxiety Disorder (GAD-7) and clinical Depression (PHQ-9) offer a baseline look of the state of student mental health in Fall 2020.^9,10,11,12,13^ Based on the scoring systems of the GAD-7 and PHQ-9, 34.8% of students were classified as having moderate to severe Generalized Anxiety Disorder, while 13.1% were classified as having moderate to severe Depression. While we cannot discern whether these rates are different from pre-pandemic rates, it is useful for comparisons across other universities and within MU over time. Additionally, these rates highlight a need for expanded mental health resources on campus.

### 3.6 How Did Self-Enrolled Students Differ from Randomly Selected Students?

The 89 students who self-enrolled into the study, without invitation, were different in important ways from those who were contacted via random selection and then chose to enroll in the study. Chi-square tests revealed that the self-enrolled were more likely to have had a previous COVID-19 PCR test, test positive for SARS-CoV-2 antibodies, be older, and be graduate students.

## 4 Conclusion

The results of our study indicate that between 11% to 15% of students had been infected with COVID-19. Further, students were less likely to report they had a chance of having COVID-19 than of knowing someone who had contracted COVID-19. The vaccine intention rate suggests herd immunity may be possible, although low rates of flu vaccine take-up suggest converting intention to actual vaccination take-up may prove challenging. Finally, student mental health concerns, especially anxiety and depression, are noted.

Our study raises several issues. We are concerned about student distrust of how the university will use their COVID health information. For instance, more than half of students were unwilling to suggest their chances of having been infected with COVID-19. In Fall 2020, students were quarantined when they reported positive test results from whatever source of testing (e.g., campus testing service, county health department). Members of the campus community suspected students did not want to be quarantined, and therefore may not have been tested even when they thought they were exposed to the virus. Students may have been unwilling to note a high chance of a being infected out of fear that the study personnel would report the information to those on campus involved with the quarantine process.

Although a high share of students indicated an intention to get the COVID-19 vaccine when available, in the months since this study there has been an onslaught of misinformation about the vaccine. Vaccine hesitancy among students may have increased. The university will need to institute public health communication strategies that encourage vaccination, possibly using advertising that reminds students that the majority of them intended to get the vaccine.

Finally, we note that men are underrepresented in our study. Given the national evidence that men are less likely to support COVID-19 mitigation strategies, and that subsets of men are most vaccine hesitant, our estimates of COVID-19 positivity rates and vaccine intentions may not be as accurate for male students, a subject of future research.^14^ Further, public health research on college students likely needs specific marketing strategies to increase participation among male students.

Keeping the student body and surrounding community safe, healthy, and informed, to the greatest extent possible, should always be a high institutional priority. This study was one strategy used by our university to address health concerns of our community. The relatively low positivity rate and high vaccine intention rate suggest, still, that public health communications on the campus were effective.

## Data Availability

Data produced in the present study are available upon reasonable request to the authors.

## 5 Acknowledgements

In Fall 2020, MU CARES funding and other internal funding sources, as well as support from SIEMENS Healthineers, funded the MU Study of Seropositivity and Risk for SARS-CoV-2 and COVID-19. We would like to thank Mark McIntosh, Stevan Whitt, John Middleton and the MU Incident Command Team who foresaw the importance of this study and facilitated its creation. We also could not have successfully collected these data without the support of Student Information Services and Human Resources to supply randomly selected email lists (Chase Hickman, Allen Johanning, Jeremy Wiebold), IT staff who helped with appointment and website development (Lei Jiang), the team of consenters (Brady Chung, Emma Foust, Emily Perisho, Kamilla Sarvestani, Peyton Kusgen, Megan Polniak, Sarah Simmons, Tess Willemse), the clinical staff (Allison Thomas, Collin Welch, Ella Bochenski, Madison Enright, Madison Hassler, Tess Goldenberger), our Student Health partners (Jamie Shutter, Scott Henderson), the REDCap survey and data collection design team (Abu Mosa, Vasanthi Mandhadi), the risk survey and qualitative study teams (Ifeolu David, Michelle Teti, Yerina Ranjit, Haejung Shin) and the immunology lab team (Alexander Earhart, Kim Laffey, Adam Schrum). Finally, we would also like to thank the students, staff and faculty in the MU community who participated in this study.

